# Physical activity and Retirement: Original analysis of responses to the English Adult Active Lives Survey 2016/17

**DOI:** 10.1101/2020.03.02.20029967

**Authors:** Julii Brainard, Rachel Cooke, Kathleen Lane, Charlotte Salter

**Author notes:** **Corresponding author:** Dr. Julii Brainard, Postal Address as above.

## Abstract

**Objectives:** Opportunities for older adults to do physical activity may depend on other commitments. To see if reported physical activity was higher or lower among older adults depending on work status: full time, part-time or retired.

**Methods:** The Active Lives Survey 2016/17 in England was used to see how active people were depending on employment or retirement status. Types of physical activity (PA) considered were: leisure, gardening, active travel and combined total, adjusted for age, sex, BMI, disability, rurality and deprivation in models using hurdle negative binomial regression.

**Results:** Total PA was significantly greater for retired persons compared to both full- and part-time workers age 55-64, while being retired or working part-time at age 65-74 meant more PA. People did more leisure or gardening with less work, but active travel decreased with fewer work hours, at all ages. Retirement meant more leisure and gardening PA but less active travel.

**Conclusions:** Demand for opportunities to engage in leisure and gardening PA appears to be high among retired people. Greater promotion of active travel in this cohort may be possible.

## Introduction

Retirement is a complex process and a major life and employment transition that impacts all aspects of health and well-being, including physical activity. Staying physically active is widely promoted to ensure good health in later years of life, yet physical activity (PA) tends to decline following retirement, especially in lower socioeconomic groups (Lloyd 2011, Yorston *et al*. 2012). Loss of occupational and travel-linked PA contribute to net reductions in PA after retirement (Berger *et al*. 2005), even as recreational and household PA tend to increase, at least in early years after retirement (Barnett *et al*. 2014).

The process of transitioning to retirement in England has been estimated to take an average 10 years (Banks *et al*. 2016); this long period should offer many types of opportunities for interventions that can compensate for reduced occupational and active travel PA that was linked to work. Understanding the perspectives of older adults about PA could also inform interventions for them. Compared to younger adults, older adults may be more aware of potential health benefits from staying active (Caudroit *et al*. 2011). Equally, retired people have distinctive perceptions of their time and energy availability for doing physical activity (McDonald *et al*. 2015, Devereux-Fitzgerald *et al*. 2018). Self-efficacy (Caudroit *et al*. 2011) and identity issues are pertinent; older adults without a past history of being physically active may find it especially difficult to envision themselves as someone who could start to routinely undertake PA (Kosteli *et al*. 2016).

As part of a wider study looking for intervention opportunities to support PA during the transition period to retirement, we were given unique access to a large and recent survey of physical activity for adults living in England. The data have not been subject to in-depth analysis previously. The survey included questions about many demographic traits, including employment status. Our primary objective was to explore if and how participation in or levels of physical activity seemed to be higher among those in work or who were retired.

## Methods

### Active Lives Survey

The Adult Active Lives Survey 2016/2017 (ALS1617) was conducted by Ipsos MORI on behalf of Sport England (Sport England 2015, Ipsos Mori 2018, Sport England 2018). Sport England is a semi-autonomous, publicly funded body tasked to promote and develop public sport and physical recreation in England, UK (Sport England 2009). A target of 500 returns was set from each local authority in England, with survey invitations sent to randomly selected addresses from a database for all United Kingdom residents maintained by the Royal Mail (encompassing all local government areas). Response rate for 2016-17 is not published but the response rate for the Active Lives Survey undertaken in 2015-16 was 18.9% (Ipsos Mori 2017). Data were collected from November 2016 to November 2017 using both web survey forms (52%) and paper questionnaires (48%). The sampling strategy was designed to be representative of the population across key demographic variables (such as gender, age, geographic spread and levels of deprivation). The sampling frame and targets were intended to elicit responses from diverse demographic and geographic areas rather than calculated to satisfy any specific statistical query. Participants were informed that their replies would be used to help provide better services. Ethics approval for this secondary analysis was not required because consent was implied by submitting the completed questionnaire. Respondents were rewarded with a £5 shopping voucher. The dataset described 198,911 individuals (age 16+), of whom 93,509 were persons age 55+ years. Although we had access to the ALS1617 we are not authorised to share the original onwards so only summary results are made available here.

The questions asked about specific physical activities people did in the preceding 28 days, duration, frequency, and whether the PA raised their breathing rate or made them sweaty. PA done for leisure or sport, gardening and active travel (cycling or walking for transport) was asked about. The questionnaire did not ask about physical activity connected to home maintenance, housework or occupation (except when occupational PA could also be categorised as active travel).

Reported PA was further categorised as moderate or vigorous by either (by respondents or assumed by questionnaire coding rules), as:

- Moderate activity: Heart rate raised to put individual a little out of breath.
- Vigorous activity: Breathing hard and fast and heart rate increased significantly

Automatic coding by the questionnaire for some types of activity into moderate or vigorous helped to reduce question burden on respondents and helped ensure consistency of categorization across the respondent group for similar activities; for instance, all walking was assumed to be moderate and all running was assumed to be vigorous. “Moderate intensity equivalent minutes” (MIEMS) were calculated for each respondent by the data provider. MIEMs have been validated as acceptably robust but not data-demanding indicators of total physical activity in population surveys (Milton *et al*. 2017). MIEMS in the ALS1617 were determined both by self-reported intensity (whether breathing rate was raised slightly or strongly) and type of activity. When calculating MIEMs, each ‘moderate’ minute counted as one minute, but a vigorous activity counted for double. For instance, a single 10-minute walk was 10 MIEMs, while a vigorous 10-minute run equalled 20 MIEMs. MIEMs were calculated from all PA sessions of at least 10 minutes’ duration, reported during the previous 28 days divided by four to produce a typical average over 7 days.

The ALS1617 also asked for gender, age, working status, disability, occupational group, educational qualifications, height and weight. Disability was defined as an individual reporting that they had a physical or mental condition that has lasted or will last at least 12 months, and that substantially affected their ability to do normal daily activities. Respondents’ residence area was categorised by deprivation level by the data provider (Sport England) and (categorised by decile within the Index of Multiple Deprivation 2015; Dept. for Communities and Local Government 2015). Each decile categorises an exclusive 10% of the entire population in England according to weighted scoring in seven social domains: employment, health, income, education, crime, barriers to services and living environment. Different groupings of the deprivation indicators were tried (alternative results not fully elucidated here). A simple two-tier distinction: three highest deciles or seven lowest deciles had best fit in the final models. The survey was also provided with an indicator of relative urban density or rurality for each respondent, using a schema developed for the Office of National Statistics (Bibby & Brindley 2012). Retaining the full range of urban/rurality categories led to the best model fits.

### Analysis of the ALS1617

We focused on the period closest to retirement for most people. Although the timing is very individual, *most* people living in England retire close to the statutory pension age (SPA Hofaecker *et al*. 2016). 65 years and 62 years were the SPAs for men and women respectively in 2016-17, with SPA rising to 65 years for women by November 2018. We found that the percentages of persons in retirement significantly rose around age 63-65 years so we stratified the data into two age bands (55-64 and 65-74 years). Within these groups, we considered all persons in full-time work, part-time work or who were fully retired. Of the 93,509 respondents who were age 55+, 74,188 were age 55-74. We did not analyse persons in some work-status categories (unemployed, students, keeping house or never worked) due to small numbers in each group and so that we could focus on differences between working and retired persons. For PA indicators, we used four MIEMs measures derived from or provided with the ALS1617: leisure PA (defined as all PA done for fun, fitness or sport, but excluding gardening and active travel), gardening PA, active travel PA, and totals of all three previous, which for brevity we call ‘total PA’. We acknowledge that our label ‘total PA’ is imperfect due to categories of PA (occupational and housework) not asked about.

For all PA indicators, the distribution of MIEMs values was skewed: mostly relatively low values (including many zeros; 19% of people age 55-74 reported zero MIEMs) with a tiny percentage of extremely high values. We applied hurdle regression, which modelled PA participation in two separate models: one model for participation in PA or not (dichotomous outcome in a logit model) and a separate model (continuous response variable) to predict amount of MIEMS among those who reported any PA (using a zero-truncated negative binomial model). The models adjusted for age, sex, BMI group, disability, season, rurality and deprivation, looking at all four types of PA. We did not include occupational group or education levels in the models due to missing data (22% for education), limited category breakdown, collinearity problems with each other and the deprivation indicator, as well as risk of informant-biases or errors (e.g., respondents had to decide their occupational group from a short list of exemplars). The ALS1617 dataset had been cleaned but still retained any plausible answers. Relatively extreme reports for MIEM values (2.0% of total), which were defined as MIEMs ≥ 3360 (equivalent to ≥ 8 hours of moderate activity, 7 days/week) were excluded to get better statistical model fit for the vast majority of observations. Tables 1 and 2 describe the independent variables used in the models and participant characteristics. Most data were available for most respondents.

**Table 1.** Variables used in regression models

| **Attribute** | **Description, with reference category indicated, where relevant** |
| --- | --- |
| Age | In whole years |
| BMI Group | Calculated from self-reported height and weight.<br>Reduced to 3 useable categories =<br>Healthy weight (reference); Underweight or overweight (both one category away from being a healthy weight); Obese or morbidly obese |
| Disability | 2 useable categories = with or without limiting disability |
| Gender | 2 useable categories = male or female |
| Index of multiple deprivation 2015 | 2 useable categories, least deprived 7 deciles as reference, variant= three most deprived deciles (ideally three deciles would have been 30% of total responses but in reality was ~ 24.6% of responses). |
| Rural/Urban classification of home address, from ONS RUCLAD data | 6 useable categories available. Categories =<br>Urban major conurbation ; Urban minor conurbation<br>Urban city and town ; Rural town ; village ; hamlet |
| Season (Quarter) when survey was submitted | Winter (16 Nov-15 Feb) was used as reference category, others =<br>Spring (16 February-15 May), Summer (16 May-15 August) and<br>Autumn (16 August-15 November) |
| Working status | Models only consider 3 categories: Working full-time (reference), working part-time or retired. Respondents were asked to select their 'main status', and defined for selves what (how many hours) FT or PT meant. |

**Table 2.**
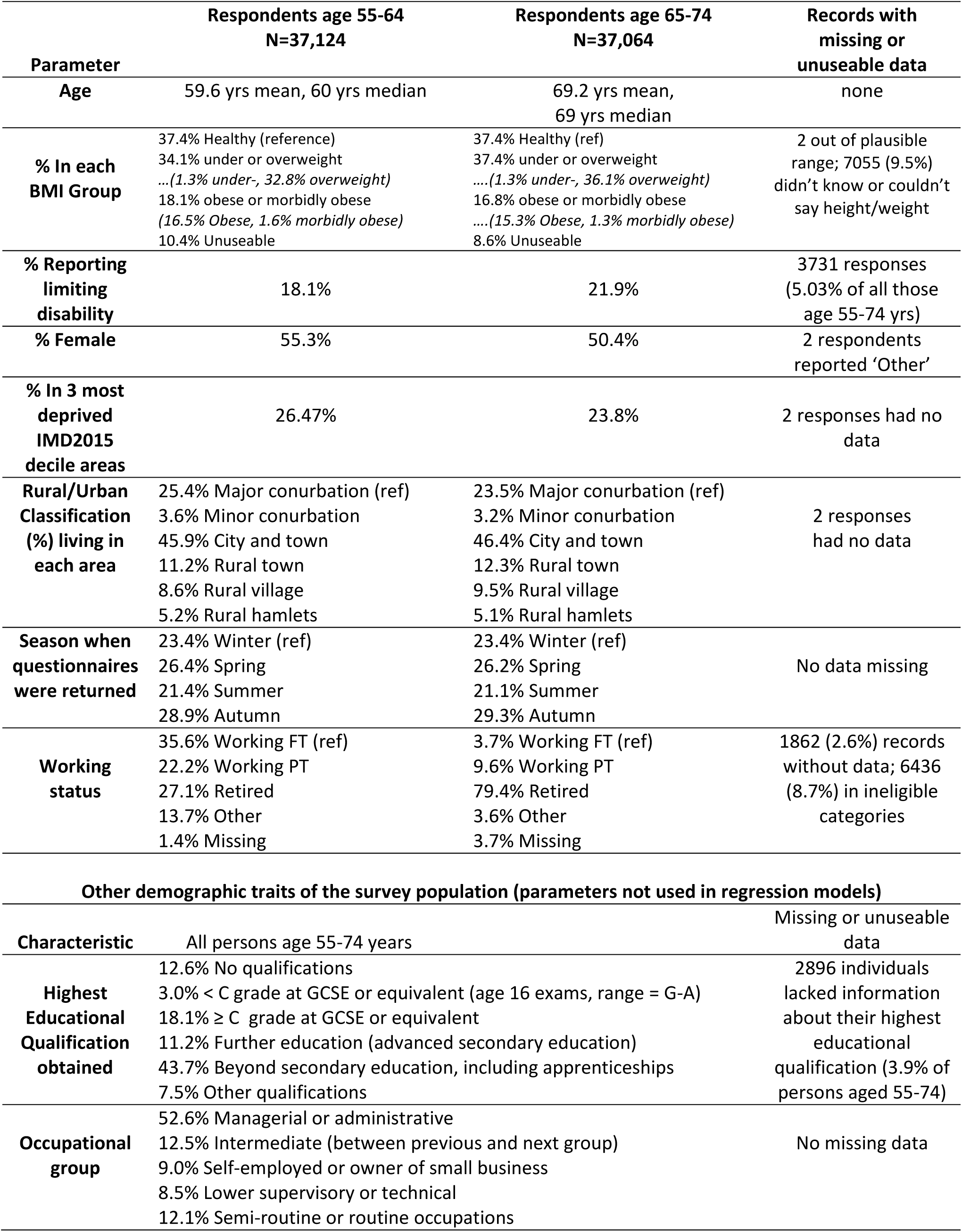
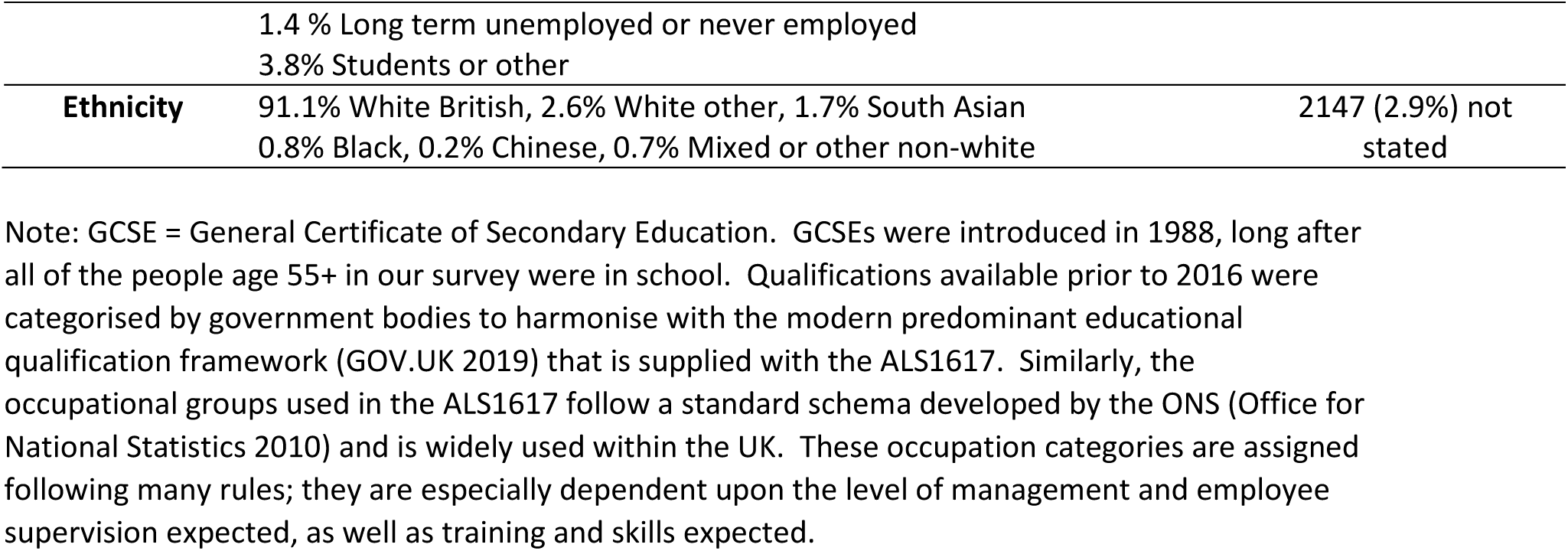
Characteristics of survey respondents.

The models treated full-time workers as the reference category. Differences between part-time workers and retired people were reported using odds ratios (any reported participation in PA model) or incidence risk ratios (MIEMs values among those who reported any PA). Data and statistical analysis were undertaken in SPSS (v. 25), MS-Excel 2016 and Stata (v. 15.1).

## Results

Full model specifications are in Supplmentary File 2 (Models 1-16), while this report focuses on work status alone. Table 3 shows the results from first stage of each hurdle model that related participation in each type of PA with work status. In adjusted models for both age groups, people tend to be more likely to report doing some leisure PA or gardening when they report less employment. With respect to leisure PA and with full-time workers under 65 as referent, part-time workers had OR 1.23 (95%CI 1.13-1.33) for leisure PA and retired persons had OR 1.48 (95%CI 1.36-1.60). Similarly, with respect to gardening MIEMS and with full-time workers under 65 as referent, part-time workers had OR 1.17 (95%CI 1.10-1.25) while likelihood of retired persons engaging in gardening had OR 1.35 (95%CI 1.26-1.44).

**Table 3.** Hurdle modelling, stage 1 (logit regression) odds ratios for participation or not in PA (in preceding 28 days)

| **Adults age 55-64** |  |  |  |  |  |
| --- | --- | --- | --- | --- | --- |
| **WORK STATUS** | Max N | All PA | Leisure PA | Gardening only | Active travel only |
| Work FT | 13,223 | 1.0 (ref) | 1.0 (ref) | 1.0 (ref) | 1.0 (ref) |
| Work PT | 8,239 | 1.26 (1.16-1.38) | 1.23 (1.13-1.33) | 1.17 (1.10-1.25) | 1.04 (0.98-1.11) |
| Retired | 10,073 | 1.54 (1.41-1.69) | 1.48 (1.36-1.60) | 1.35 (1.26-1.44) | 0.91 (0.85-0.97) |
| **Adults age 65-74** |  |  |  |  |  |
| **WORK STATUS** | Max N | All PA | Leisure PA | Gardening only | Active travel only |
| Work FT | 1360 | 1.0 (ref) | 1.0 (ref) | 1.0 (ref) | 1.0 (ref) |
| Work PT | 3553 | 1.65 (1.40-1.95) | 1.71 (1.46-1.99) | 1.26 (1.09-1.45) | 1.34 (1.16-1.56) |
| Retired | 29,412 | 1.60 (1.39-1.85) | 1.61 (1.41-1.83) | 1.37 (1.20-1.55) | 0.94 (0.82-1.07) |

**Notes:** OR = odds ratios (adjusted). N refers to maximum possible rather than actual number of observations for each model (missing data meant actual numbers were lower). 95% confidence intervals for stated OR are in (). All OR reported in Table 3 are significant at $p \leq 0.05$ . FT= full-time (working), PT= part-time. IRR = incidence risk ratio. All models adjust for age, sex, presence of limiting disability, seasonal quarter, deprivation, BMI and urbanness/rurality of residence.

Among the under-65s, propensity to engage in active travel was similar for FT and PT workers. FT workers were the referent and OR for part-timers was insignificantly different with OR 1.04 (95%CI 0.98-1.11). However, retired persons under 65 were much less likely to engage in active travel, OR 0.91 (95%CI 0.85-0.97).

In contrast, any participation in active travel was more common for age 65-74 part-time workers (OR 1.34, 95%CI 1.16-1.56) than same age group full-time workers (referent) *or* retired persons (OR 0.94, 95%CI 0.82-1.07). Among respondents age 65-74, there was insignificant difference in likelihood of participation in active travel between full-time workers and the retired. This last result could arise from the relatively small number of persons in full-time employment in the age 65-74 group (n=1360). Among the age 65-74 respondents, both retired persons (OR 1.71, 95%CI 1.46-1.99) and part-timers (OR 1.61, 95%CI 1.41-1.83) reported significantly more propensity to undertake leisure PA than did the referent full time workers. Gardening was similarly more likely among the part-timers and retired than among people working full time.

Table 4 shows the relationship between work status and median MIEMs/week in each PA category, among those who engaged at all in each type of PA (age stratified). Differences are reported as incidence risk ratios (IRR) with 95% confidence intervals. IRR with 95% confidence intervals entirely below 1.0 strongly suggested *less* active-travel PA for retired persons but IRR with 95% confidence intervals above 1.0 suggest *higher* leisure and gardening PA for retired persons.

**Table 4.** Stage 2 hurdle models (zero-truncated negative binomial). Dependent variable = MIEMs, amount of activity undertaken, for those who reported participating in PA at all. Incidence risk ratios (IRR) relative to working full time, for four categories of physical activity, adults age 55-74

| ADULTS AGE 55-64 |  |  |  |  |  | ADULTS AGE 65-74 |  |  |  |  |
| --- | --- | --- | --- | --- | --- | --- | --- | --- | --- | --- |
| WORK STATUS | Max N | median MIEMs | Difference from working FT | adjusted IRR | Adj. IRR 95%CI | N | median MIEMs | Difference from working FT | adjusted IRR | Adj. IRR 95%CI |
| **ALL PHYSICAL ACTIVITY** |  |  |  |  |  |  |  |  |  |  |
| Work FT | 10,594 | 510 | - | 1.0 (ref) | - | 979 | 480 | - | 1.0 (ref) | - |
| Work PT | 6798 | 530 | +20 | 1.05 | 1.02-1.08 | 2833 | 500 | +20 | 1.13 | 1.05-1.21 |
| Retired | 8329 | 645 | +135 | 1.23 | 1.20-1.27 | 22,482 | 510 | +30 | 1.21 | 1.13-1.28 |
| **LEISURE ONLY, WHICH EXCLUDES GARDENING AND ACTIVE TRAVEL** |  |  |  |  |  |  |  |  |  |  |
| Work FT | 10,127 | 435 | - | 1.0 (ref) | - | 899 | 380 | - | 1.0 (ref) | - |
| Work PT | 6556 | 450 | +15 | 1.04 | 1.01-1.08 | 2700 | 407 | +27 | 1.10 | 1.02-1.19 |
| Retired | 8056 | 540 | +105 | 1.21 | 1.17-1.25 | 21,025 | 420 | +40 | 1.18 | 1.10-1.26 |
| **GARDENING ONLY** |  |  |  |  |  |  |  |  |  |  |
| Work FT | 4224 | 180 | - | 1.0 (ref) | - | 426 | 240 | - | 1.0 (ref) | - |
| Work PT | 2742 | 180 | 0 | 1.06 | 1.00-1.12 | 1240 | 240 | 0 | 1.08 | 0.96-1.21 |
| Retired | 3744 | 240 | +60 | 1.27 | 1.21-1.34 | 10,436 | 240 | 0 | 1.19 | 1.07-1.32 |
| **ACTIVE TRAVEL ONLY** |  |  |  |  |  |  |  |  |  |  |
| Work FT | 4642 | 180 | - | 1.0 (ref) | - | 379 | 180 | - | 1.0 (ref) | - |
| Work PT | 2898 | 150 | -30 | 0.81 | 0.77-0.86 | 1156 | 150 | -30 | 0.83 | 0.72-0.95 |
| Retired | 3155 | 148 | -32 | 0.82 | 0.77-0.87 | 7199 | 150 | -30 | 0.80 | 0.71-0.90 |

**Notes:** MIEMs calculated as described in text (minutes of moderate intensity exercise, over 7 days), among only those who reported some PA. “Difference from working FT” refers to difference in median MIEMs. FT= full-time (working), PT= part-time. IRR = incidence risk ratio. All models adjust for age, sex, presence of limiting disability, seasonal quarter, deprivation, BMI and urbanness/rurality of residence. Work status was significant at $p \leq 0.05$ in all models.

### Physical activity: Total and Leisure

Leisure was the main type of activity generating MIEMs for most people and dominate the aggregate results. Full-time workers age 55-64 reported significantly less total or leisure PA than people working part-time. For leisure PA, with full-time workers as referent, part-time workers had IRR 1.04 (95%CI 1.01-1.08) and the corresponding IRR for retired people was 1.21 (95%CI 1.17-1.25, p<0.001). At age 65-74, the also retired reported more leisure PA than workers. Retired and part-timer median reported MIEMs were respectively 407 and 420, which did not seem to be significantly different from each other (evidenced by overlapping IRR confidence intervals) for age 65-74. Both retired (IRR 1.18, 95%CI 1.10-1.26) and part-time workers (IRR 1.10, 95%CI 1.02-1.19) age 65-74 reported significantly more leisure MIEMs than full time workers.

### Gardening

People were much more likely to report doing any gardening if retired or part-time employed than if working full-time. The median reported MIEMS spent gardening was relatively consistent, either 180 MIEMS (age 55-64 working PT or FT) or 240 MIEMS (retired persons age 55-64 and all persons age 65-74). Nevertheless, significantly more gardening MIEMs were done by age 55-64 part-timers (IRR 1.06, 95%CI 1.00-1.12), age 55-64 retired persons (IRR 1.27, 95%CI 1.21-1.34) and age 65-74 retired persons (IRR 1.19, 95%CI 1.07-1.32). It should be stated that gardening was still a minority past-time; 67% of people age 55-74 reported no gardening in the preceding four weeks (so recorded zero MIEMs). Among those who did any gardening PA, gardening comprised (on average) about 45% of total reported MIEMs whether working or retired.

### Active Travel

For the age 55-64 group, reported levels of active travel in Table 4 were significantly lower in those who were part-time and retired, compared to those working full-time (median 150 or 148 MIEMs vs. 180 MIEMs, approximate IRR 0.81, p<0.001). At age 65-74 years, there was also significant difference in active travel participation between full-time workers and either part-timers or the retired (median 180 MIEMs vs. 150 MIEMs, approximate IRR 0.83). An alternative comparison for impacts of work status on active travel PA may provide better insight to whether more work seems to encourage active transport. Instead of comparing MIEMS for the PT or retired to FT workers, when we compare FT vs. combined group of PT+retired at age 65-74, this yields OR = 0.82 (95%CI 0.71-0.93). 67% of people age 55-64 and 75% of those age 65-74 reported no active travel (walking or cycling) in the preceding four weeks. For those who did any active travel, active travel comprised (on average) approximately 37% of reported MIEMs, whether working or retired.

## Discussion

Reported leisure- and gardening-related physical activity among persons age 55-74 was greatest when retired, and much greater than reported by full-time workers. Reported leisure and gardening PA among retired persons age 55-64 was also greater than for part-time workers of the same age, but this difference for part-time workers and retired people was negligible for age 65-74. The reported increase in leisure and gardening PA was greater than reported decline in active travel for the same comparator groups.

There are many potential “favourable and unfavourable lifestyle changes” at retirement that can influence total PA (Zantinge *et al*. 2013). These changes impact maintenance, sustainability of feasible forms of PA, motivations, financial resources, personal circumstances (such as caring responsibilities), perceived benefits of PA, resilience and social expectations. Some longitudinal studies found that retirees reported significantly greater PA, particularly in walking and moderate-intensity activities, compared with pre-retirement. From longitudinal self-reported PA data (Barnett *et al*. 2014) in models adjusted for age and multiple other attributes, leisure and household PA seem to generally rise after retirement even as active travel decreased within an English population. However, Ding et al. (Ding *et al*. 2016) observed that the “activity-promoting effect” of retirement is likely to most benefit those who retired at a younger age, who have better baseline physical function, and/or those who worked full-time prior to retirement. Evidence on other populations also suggested that while leisure-time PA tends to increase among the retired and those transitioning to retirement, overall PA does not necessarily increase (Hobbs *et al*. 2013, McDonald *et al*. 2015) and net total PA may in fact decrease post-retirement (Holstila *et al*. 2017). In longitudinal analysis, Stenholm et al. (2016) observed an early sharp rise in physical activity in the first few years after retirement, followed by decline to pre-retirement levels typically within 5-10 years. Vigorous PA levels had a linear decline in older adults with increased age that was unaffected by retirement.

Increased age alone means increased risk of poor health or disability that can make PA more difficult (Franco *et al*. 2015, Büchs *et al*. 2018). Socioeconomic status and gender are important factors that can interact with quality and quantity of physical activity throughout the life course, including among older adults (Barnett *et al*. 2012, Barnett *et al*. 2013). Participation barriers identified for older adults include lack of confidence, apathy, and lack of appropriate activities or activity leaders (Franco *et al*. 2015). Older adults are highly influenced by environmental features when deciding whether to engage in outdoor PA. Unpleasant neighbourhood features (such as litter or lack of pedestrian paths) are discouraging, while attractive environmental features (such as parks and cafes) seem to encourage greater PA (Franco *et al*. 2015, Cerin *et al*. 2017).

Separate from the effects of increased age, retirement impacts PA in other ways. There may be more time for physical activity, but also more potential for competing activities that are higher priority, such as caring responsibilities (Franco *et al*. 2015, International Longevity Centre 2017). Reduced income is a possible barrier (Franco *et al*. 2015), as well as the loss of a daily structure which previously enabled and facilitated PA (McDonald *et al*. 2015, Banks *et al*. 2016, Kosteli *et al*. 2016).

Decline in active travel following retirement is posited to relate to loss of structure and routine that were provided by previous occupational duties. One way that structure (that facilitates PA) could be regained is via activities like dog-walking, gardening and regular voluntary work activities. Voluntary work examples are conservation, leading walking groups or sports coaching, which have the potential to beneficially replace physical activity opportunities that arose due to employment activity. A policy in Britain known to successfully increase active travel (walking) is provision of free local bus passes for older persons (Coronini-Cronberg *et al*. 2012). Perhaps in contrast to active travel, gardening is a type of PA socially acceptable to older adults and that conforms with identity expectations about social position and advancing age (Bhatti 2006). Determinants and motivators for doing PA are often described as highly individual (McDonald *et al*. 2015), and the best theoretical framework for designing physical activity interventions that target older people or adults in transition to retirement remains unclear (Morgan & Tan 2018).

Because the ALS1617 data are cross-sectional, we cannot confirm *change* in activity after retirement or due to retirement. However, the implications are clear: retired people report more leisure and gardening PA, but less active travel PA, than working persons of the same age. A picture also emerges of a minority of very active older adults who several times over met the UK Chief Medical Officers’ (CMO) guidelines to achieve at least 150 MIEMs/week (Chief Medical Officers 2011). Of those respondents (retired or still working) who engaged in any active travel at least 50% met the CMO guidelines from active travel alone. The same is true of respondents who engaged in any gardening; at least 50% met the CMO guidelines from gardening alone. Many older adult respondents to the ALS1617 demonstrated ample appetite to undertake PA during retirement, at least within the leisure and gardening PA categories. We have also evidenced a widespread belief (but not often documented in scientific literature) that older adults like gardening; gardening was the second most popular physical activity in the previous year for ALS1617 respondents age 55+ (Supplementary file 1, Tables 1-2).

### Limitations

Our analysis could not address differences in housework or occupational PA (as this was a secondary data analysis and that information was not collected in the original survey), and hence we could not evaluate subsequent potential impact on either total true PA or health outcomes. Note that estimating occupational PA before retirement is difficult because often older workers move out of roles that have high occupational PA into short- or medium-term (“bridge”) occupations that are less physically demanding (Johnson *et al*. 2009). It merits mention, too, that the health benefits of occupational PA are contested (Holtermann *et al*. 2012, Coenen *et al*. 2018).

To focus on the specific possible effects of retirement we excluded many work status categories: unemployed, students, having never worked or long-term unable to work due to sickness/disability; we have no findings about these other populations. We categorised and stratified the dataset to make interpretation more meaningful and associations more apparent; different categorisation schema would have led to somewhat different raw incidence risk and odds ratios, but we don’t believe those variations would substantially change the main conclusions or associations that we observed.

The ALS1617 data were self-reported and therefore prone to recall, subgrouping and engagement biases. Generalisability of our observations is also limited due to imperfect representativeness of English residents age 55+. Within the ALS1617 data, the percentages of age 55-74 persons still in employment, living in not deprived areas, in administrative or managerial occupations or with healthy BMIs were greater than observed nationally (Office for National Statistics 2016, Baker 2018). ALS respondents also report more PA than the general population. In the 2016 Health Survey for England (NHS Digital 2017), about 55% of respondents age 55-74 reported obtaining ≥ 150 minutes of PA per week, compared to 66% of same-age ALS respondents who reported reaching this threshold.

## Conclusions

Retired people reported doing more leisure and gardening PA but less active travel. Some older adults reported enough physical activity from either gardening or active travel alone to meet official recommendations for best health outcomes. People working full time reported less leisure PA and less gardening PA than people with retired status, adjusted by age. Policies to promote recommend amounts of regular physical activity for older adults need to acknowledge different opportunities that may be facilitated by working status.

## Data Availability

We do not have authority to share the original data which are owned by Sport England. There is an aspiration to put these data on UK Data Archive in future.

## Acknowledgements

Thanks to James Goldson of Sport England (SE) for supplying us with the survey data and answering many questions about it. Nicola Wildash of SE gave helpful comments on our draft manuscript. Paul Hunter (UEA) got us started with hurdle regression. We are also grateful to Andy Jones and Karen Milton of UEA who shaped our protocol and study objectives.

## Supplementary Information

Supplementary File 1, Table 1. Ten most popular physical activities in previous year, from ALS1617 survey (women)

Supplementary File 1, Table 2. Ten most popular physical activities in previous year, from ALS1617 survey (men)

Supplementary File 2, Models 1-16. Logit (participation in each type of PA) models and amount of MIEMS reported by those who participated at all (zero truncated negative binomial regression).

